# Genome-wide association study identifies novel risk variants for celiac disease in the 5p15.33 locus: insights from a population-based screening of adults, the HUNT study

**DOI:** 10.1101/2024.12.09.24318711

**Authors:** Mohammad Sayeef Alam, Laurent F. Thomas, Ben Brumpton, Kristian Hveem, Knut E. A. Lundin, Sebo Withoff, Iris H. Jonkers, Ludvig M. Sollid, Rebecka Hjort, Eivind Ness-Jensen

**Affiliations:** HUNT Center for Molecular and Clinical Epidemiology, NTNU, Norwegian University of Science and Technology, Trondheim, Norway; HUNT Research Centre, NTNU, Norwegian University of Science and Technology, Levanger, Norway; Department of Clinical and Molecular Medicine, NTNU, Norwegian University of Science and Technology, Trondheim, Norway; BioCore - Bioinformatics Core Facility, NTNU, Norwegian University of Science and Technology, Trondheim, Norway; Department of Gastroenterology, Oslo University Hospital - Rikshospitalet, Oslo, Norway; Norwegian Coeliac Disease Research Centre, Institute of Clinical Medicine, University of Oslo, Oslo, Norway; Department of Genetics, University Medical Center Groningen, University of Groningen, Groningen, Netherlands; Department of Immunology, Oslo University Hospital - Rikshospitalet, Oslo, Norway; Department of Medicine, Levanger Hospital, Nord-Trøndelag Hospital Trust, Levanger, Norway; Department of Molecular Medicine and Surgery, Karolinska Institutet & Karolinska University Hospital, Stockholm, Sweden

**Keywords:** celiac, gluten, autoimmune, GWAS, non-HLA

## Abstract

Previous studies have uncovered genetic loci associated with celiac disease (CeD) within both the human leukocyte antigen (HLA) and non-HLA regions. However, half of the heritability remains unexplained. This study aimed to identify novel loci associated with CeD in a general adult population screened for the disease, mitigating the likely selection bias observed in previous case-control studies. The study utilized data from the fourth Trøndelag Health Study (HUNT4) in Norway, where 52,358 adults were screened for CeD using serology, identifying 465 previously undiagnosed biopsy-confirmed cases. Additionally, 377 previously diagnosed cases were identified through hospital journal searches and registry data. Genotyping of 373,185 single nucleotide polymorphisms was performed on all participant using four Illumina HumanCoreExome arrays. Imputation, using the Haplotype Reference Consortium panel, resulted in approximately 24.9 million variants, post quality control. A genome-wide association study was performed using SAIGE, and functional mapping and pathway enrichment analysis was conducted using FUMA. All except one of the 42 known autosomal loci were present in the data, of which seven reached the suggestive significance threshold (P ≤ 5 × 10^−6^). Thirteen independent novel associations were observed (P ≤ 5× 10^−8^), with the 5p15.33 locus showing the highest potential for a true association with CeD, warranting further studies to validate the findings. Notably, the *IRX1* gene, located close to the 5p15.33 locus has also been associated with rheumatoid arthritis, suggesting a new shared autoimmune locus.

## Introduction

Celiac disease (CeD) is a chronic and multifactorial disease affecting one in every 100 individuals of European ancestry (1). CeD is caused by an immune response in the gut against dietary gluten proteins present in wheat, rye, and barley. The immune response leads to an enteropathy of the small intestine, regarded as the hallmark of CeD. The presentation of the enteropathy ranges from minor inflammatory changes in the small intestinal villi to total blunting of the mucosa (2).

Genetic studies in CeD have come a long way from establishing the role of human leukocyte antigen (HLA) DQ2.5, DQ2.2 and DQ8 allotypes to understanding the intricate mechanism of presenting cereal peptides to the CD4+ T-cells (3–7). Simultaneously, the global prevalence and incidence of CeD have risen, driven primarily by increased awareness and improved diagnostic technologies (8), as well as a documented increase in disease occurrence (9). Despite these advancements, screening has revealed high rates of undiagnosed cases (10,11). Although all CeD patients carry one or two of the DQ2.5, DQ2.2 and DQ8 risk allotypes, up to 55% of the general population also possess them (12). However, only 3% of the carriers develop the disease, indicating that HLA-DQ allotypes are necessary but not sufficient for CeD development. So far, the 42 loci discovered through genome-wide association studies (GWAS) using immune-based SNP arrays have only explained 48% of the genetic variation (13–16). While such studies have higher power to detect variants in the HLA and immune loci, they have lower coverage of variants outside these regions, which may contribute to the unexplained variation. Previous GWASs on CeD are also prone to information bias, as they often include only patients with a previous diagnosis, assigning undiagnosed and potential CeD cases to the control groups. Additionally, these studies are more prone to selection bias due to the lower response rate compared to the current study (17).

The aim of this study was to identify novel genetic variants associated with CeD in the non-HLA regions by overcoming biases in previous GWASs. We used a large adult population screened for CeD, including both known and previously unknown cases, and employed a SNP array and imputation methods that provide better coverage of the non-HLA regions than previous studies.

## Material and Methods

### Study population

The study was based on the fourth round of the Trøndelag Health Study (HUNT4), conducted between 2017-2019 in Nord-Trøndelag County, Norway. All individuals aged 20 years or older were invited to the survey. Out of 103,800 invitees, 30,598 women and 25,444 men participated, resulting in a 54% response rate.

The survey collected diverse types of data from questionnaires and field stations were established to conduct interviews and collect clinical measurements by trained personnel. Blood samples were also collected and stored in automated freezers at HUNT Biobank at -80°C for future analysis (18). Further details on the HUNT4 survey can be found in (19).

### Celiac disease screening

The screening for CeD was conducted in the following steps. First, all eligible serum samples from the HUNT4 participants (n=56,042) were analyzed for transglutaminase 2 (TG2) immunoglobulin (Ig) A and IgG antibodies at Oslo University Hospital utilizing a novel serological assay (20). Second, all seropositive individuals (i.e., TG2-IgA ≥0.7 mg/L or TG2-IgG ≥1 mg/L) were invited for a clinical evaluation at Levanger Hospital, Nord-Trøndelag Hospital Trust. This included endoscopic examination with small intestinal biopsies and repeated serological testing to confirm the CeD diagnosis.

Histopathological and immunohistochemical examinations were conducted at St. Olav’s Hospital, Trondheim University Hospital. Finally, the CeD diagnosis was based on stringent criteria, including repeated positive serology, a minimum Marsh grade of 3 (villous blunting) (21,22), and exclusion of other causes of inflammation and atrophy, such as the use of non-steroid anti-inflammatory drugs or acetylsalicylic acid and infection with *Helicobacter pylori*.

All individuals surpassing the serological threshold but with a Marsh grade 0-2 were termed potential cases. Seropositive cases were defined as known cases if they received a CeD diagnosis prior to the survey, and as new cases if they were diagnosed during the survey. Seronegative known cases were identified through journal searches and registry data.

Out of 52,358 participants with available genotype and phenotype information, 1,107 (2.1%) were found to be seropositive, and 826 (1.5%) had biopsy-proven CeD (Figure 1). Further details about the cohort and screening procedure can be found in (23).

**Figure 1:**
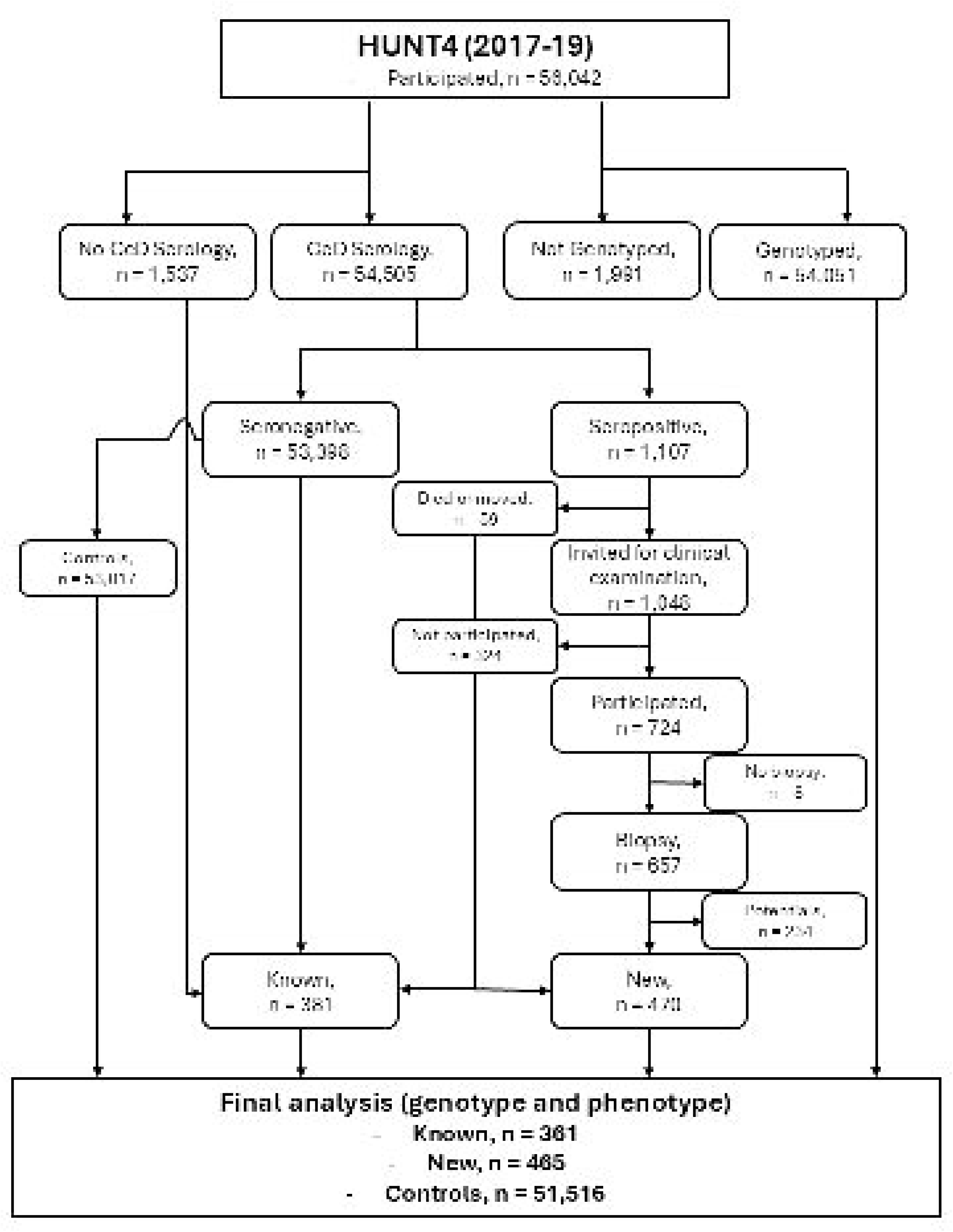
Participant Flowchart. The flowchart illustrates the inclusion and exclusion criteria for participants in the study. HUNT4=Fourth round of the Trøndelag Health Study.

### Genotyping, quality control, and imputation

Genotyping was done using four different Illumina HumanCoreExome arrays (HumanCoreExome12 v1.0, HumanCoreExome12 v1.1, UM HUNT Biobank v1.0, and UM HUNT Biobank v2.0). For quality control (QC), samples with a call rate <99% or a Hardy-Weinberg deviation with p-value <0.0001 were excluded from the dataset. Finally, 358,964 polymorphic variants passed the QC and were included. Around 24.9 million variants were imputed (imputation score >0.3) from the Haplotype Reference Consortium (HRC) panel (24) using the Positional Burrows Wheeler Transform (PBWT), a more efficient haplotype phasing method implemented in IMPUTE5 (25). For the analysis of the HLA region, approximately 12,000 variants with an imputation accuracy of at least 0.8 were included. More details about the genetics of the HUNT cohort have been described previously in (26).

### Association analyses

The SAIGE tool was employed to conduct a GWAS using a logistic mixed modelling approach (27). SAIGE was selected for its ability to account for sample relatedness and case control imbalance, as well as its efficiency in scaling to biobank level sample sizes. The phenotype of interest was biopsy confirmed CeD, with the genetic variants, sex, birth year, genotyping batch, and the first 20 principal components included as covariates.

The HLA region on chromosome 6, spanning between 29-34MB (hg19 build), was analyzed separately. A conditional logistic mixed model approach in SAIGE was implemented to determine the lead SNPs associated with CeD. The HLA lead SNP, along with all variants in linkage disequilibrium (LD, R^2^ > 0.2), and any former lead SNPs detected in this region, were iteratively conditioned on (n=3) until a non-significant lead SNP appeared.

All analysis and subsequent visualization were conducted in RStudio (28) running R v4.1.2 (29), GWASLab v3.4.45, Plink v1.9 (30) and Plink2 (31) tool in Ubuntu 22.04.6 LTS operating system. Additionally, all positions reported were based on GRCh37 build. Map2NCBI, an R package, was used to map the SNPs to the closest genes (32). The gene summaries were derived from the online GeneCards platform (33). Mapping of variants to corresponding genes was done through manual searches on dbSNP and OpenTargets.

### Functional mapping and annotation

FUMA, a web-based platform (34), was used for functional mapping by analyzing all SNPs from the GWAS, excluding the HLA region. The SNPs prioritized by FUMA were annotated and mapped to candidate genes using the SNP2GENE function, while the GENE2FUNC function explored the biological context of the mapped genes through gene-tissue heatmaps and differentially expressed gene (DEG) scores (30). LD was calculated based on the European 1000G reference panel, with remaining parameters set to default.

### Ethics approval and consent

The Regional Committee for Ethics in Medical Research Central approved the HUNT Study (#67445), the genotyping of the participants (#2014/144, #2018/1622, #152,023), and the present study (#7943). Written informed consent was received from all participants. The study was conducted in accordance with principles established by the Declaration of Helsinki.

## Results

### Non-HLA loci associated with celiac disease

The association testing of 826 confirmed CeD cases and 51,516 controls revealed 15 new genome-wide significant (P ≤ 5 × 10^−8^) SNPs in 11 novel loci (2p16.2, 2q35, 5p15.33, 6p21.2, 12p12.1, 14q11.2, 14q31.3, 15q21.1, 15q21.3, 15q21.3, 17q23.2, 18p11.23) and one known locus (2p16.1) (Table 1). Among these, eight loci had a minor allele count (MAC) > 10. Only the 5p15.33 locus had a minor allele frequency (MAF) > 5% and more than one significantly associated SNP. The remaining loci were excluded as they were likely sporadic associations. Out of the 41 previously reported non-HLA loci (13,15,35), markers from all except one were present in the current cohort with P < 0.0001. Of these, six loci (2p16.1, 2q12.1, 3q28, 6p23.3, 10p15.1, 16p13.13) attained suggestive genome-wide significance (P ≤ 5 × 10^−6^) (Figure 2). The lead variant in the 5p15.33 locus was identified as rs32727 (5:3452886_G(REF)/C(ALT); MAF = 35.82% and P ≤ 4.7 × 10^−9^). Additionally, rs32723 (5:3452578_T/C; MAF = 35.81% and P ≤ 5.93 × 10^−9^) and rs32726 (5:3451906_T/G; MAF = 35.73% and P ≤ 5.44 × 10^−9^), both in high LD with the lead SNP (>0.8), were detected (Figure 3). The findings remained consistent even after partitioning the control population into six subgroups each with 8586 controls selected without replacement. (Figure S1). The lead SNP was mapped to the *LINC01019* gene, a long non-coding RNA with the nearest protein coding gene identified as *IRX1* in dbSNP.

**Figure 2:**
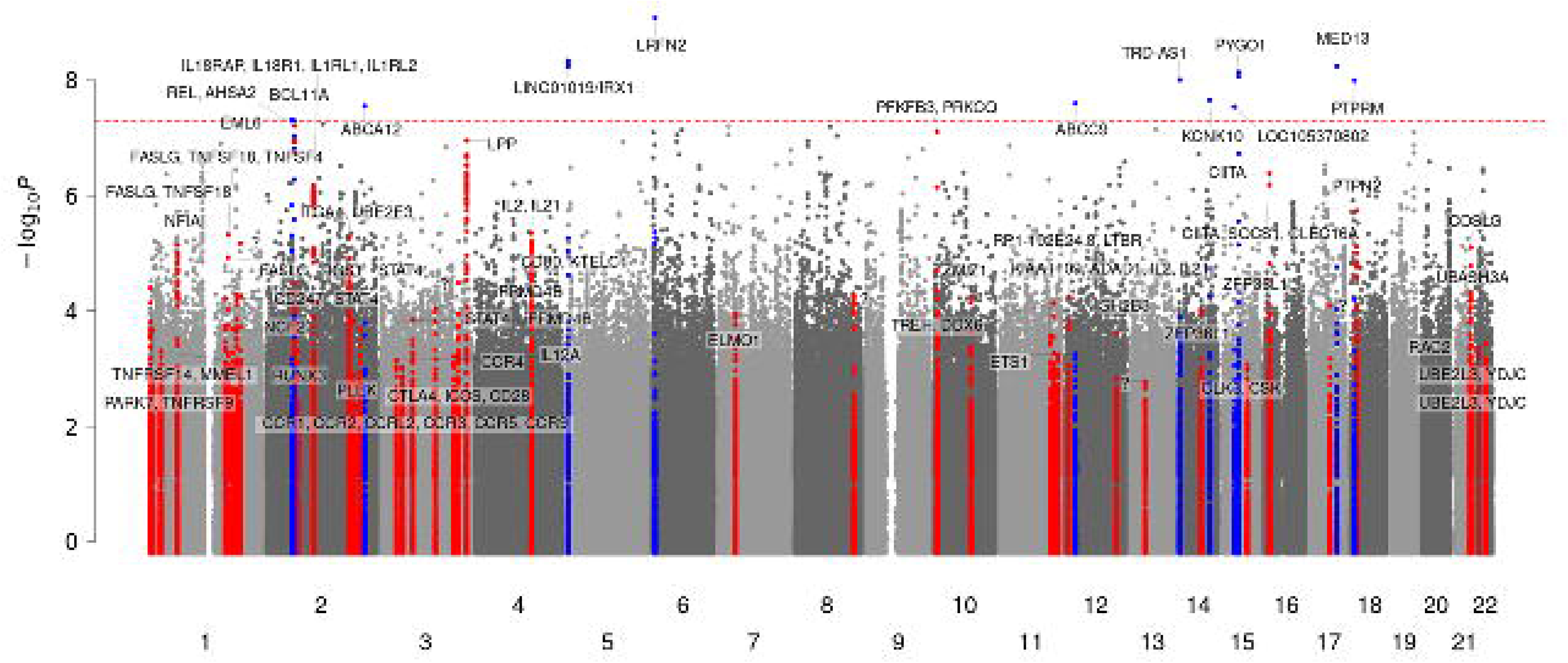
Manhattan Plot of Known and Novel Loci. Manhattan plot showing the significance levels of genetic loci (index variant ± 250 kilobase pairs) associated with confirmed celiac disease in HUNT4. Novel loci are highlighted in red, while known loci are displayed in blue. The significance threshold (P ≤ 5 × 10^−8^) is indicated by the red dotted line. Genetic variants are plotted according to chromosome and position (x-axis) and the -log_10_P for the variant association (y-axis). HUNT4=Fourth round of the Trøndelag Health Study. Sample size N=52,358. Minimum minor allele count is 3.

**Figure 3:**
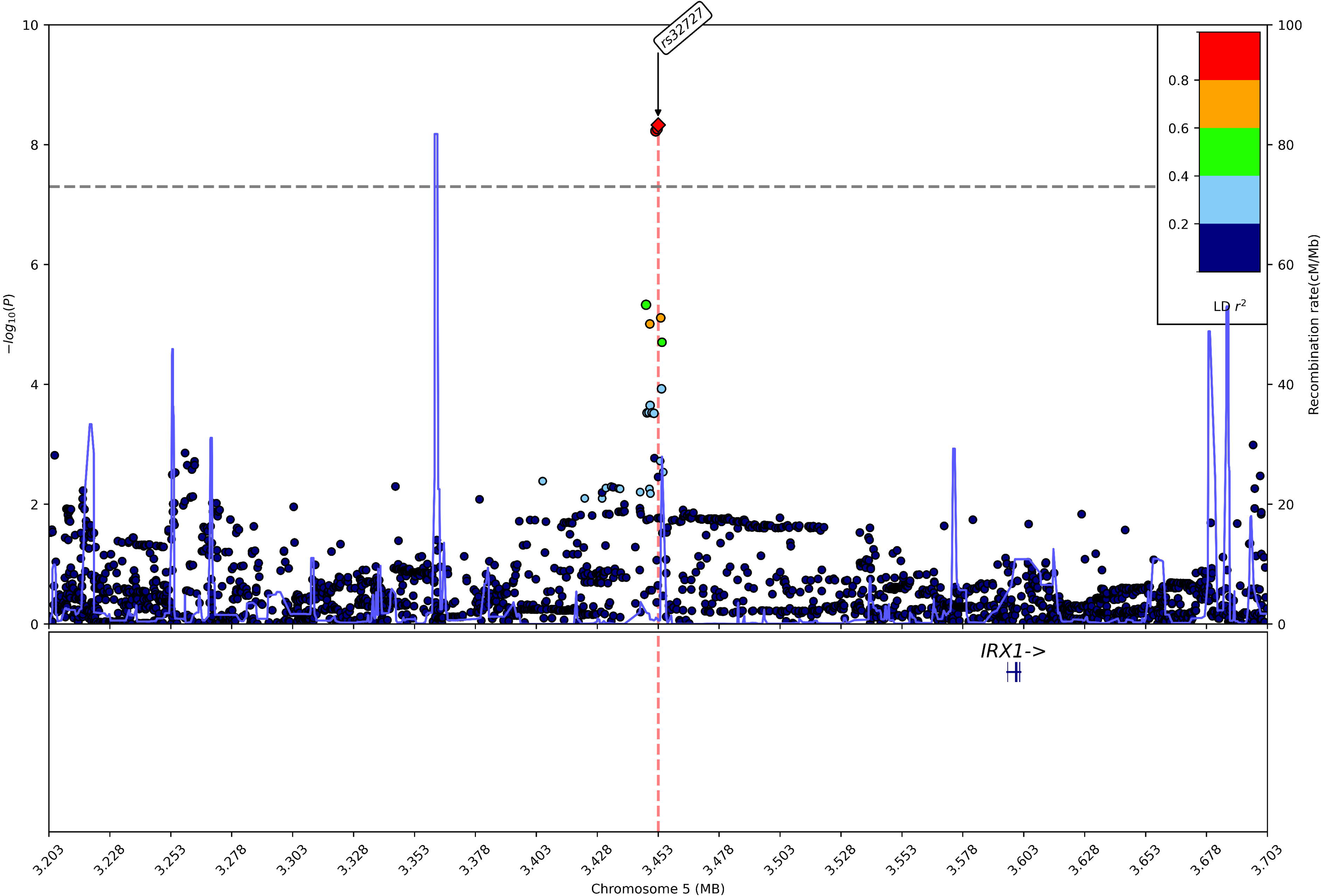
Regional Plot of the 5p15.33 Locus. Genetic variants are plotted according to chromosome and position (x-axis) and the -log_10_ P for the variant association (y-axis). The significance threshold (P ≤ 5 × 10^−8^) is indicated by the grey dashed line. The correlation r^2^ for each variant is indicated by colors relative to the index variants ± 250 kilobase pairs. The lead variant rs32727 is a long non-coding RNA with *IRX1 as the* closest protein coding gene, located 142.9 kilobases away. Two more variants, rs32726 and rs32723, were in high LD (>0.8) with the lead variant. LD=Linkage disequilibrium.

**Table 1:** Newly identified genome wide significant variants in the non-HLA-region associated with celiac disease in the HUNT4 study.

| CHR | BP | REF | ALT | ID | RSID | MAC | MAF | AF Cases | AF Controls | BETA | SE | PVALUE | Genes |
| --- | --- | --- | --- | --- | --- | --- | --- | --- | --- | --- | --- | --- | --- |
| 2 | 54974119 | C | G | 2:54974119_C/G | rs576626084 | 14.18 | 0.0001 | 0.0001 | 0.0020 | 34.5 | 6.3 | 4.67E-08 | EML6 |
| 2 | 60664486 | C | G | 2:60664486_C/G | rs551170288 | 46.65 | 0.0004 | 0.0004 | 0.0048 | 11.2 | 2.1 | 4.88E-08 | MIR4432HG, BCL11A |
| 2 | 216021220 | G | T | 2:216021220_G/T | rs189838725 | 68.14 | 0.0007 | 0.0006 | 0.0051 | 11.0 | 2.0 | 2.75E-08 | ABCA12, ATIC |
| 5 | 3451906 | T | G | 5:3451906_T/G | rs32723 | 37419.1 | 0.3573 | 0.6437 | 0.5807 | -0.3 | 0.1 | 5.93E-09 | LINC01019 |
| 5 | 3452578 | T | C | 5:3452578_T/C | rs32726 | 37500.9 | 0.3581 | 0.6429 | 0.5798 | -0.3 | 0.1 | 5.44E-09 | LINC01019 |
| 5 | 3452886 | G | C | 5:3452886_G/C | rs32727 | 37509.6 | 0.3582 | 0.6428 | 0.5794 | -0.3 | 0.1 | 4.70E-09 | LINC01019 |
| 6 | 40402692 | G | A | 6:40402692_G/A | rs192900921 | 192.37 | 0.0018 | 0.0017 | 0.0099 | 5.5 | 0.9 | 8.89E-10 | LRFN2 |
| 12 | 22005549 | T | C | 12:22005549_T/C | rs761616279 | 10.93 | 0.0001 | 0.0001 | 0.0021 | 53.8 | 9.7 | 2.48E-08 | ABCC9 |
| 14 | 22910740 | C | T | 14:22910740_C/T | rs780153546 | 3.95 | 0.0000 | 0.0000 | 0.0013 | 91.0 | 15.9 | 9.94E-09 | LOC105370401 |
| 14 | 88777584 | C | T | 14:88777584_C/T | rs769377780 | 12.76 | 0.0001 | 0.0001 | 0.0023 | 40.2 | 7.2 | 2.18E-08 | KCNK10 |
| 15 | 46555535 | G | A | 15:46555535_G/A | rs137888770 | 4.45 | 0.0000 | 0.0000 | 0.0015 | 77.8 | 14.0 | 2.84E-08 | LOC105370802, SEMA6D |
| 15 | 55560199 | C | T | 15:55560199_C/T | rs1002929661 | 5.95 | 0.0001 | 0.0000 | 0.0024 | 62.8 | 10.9 | 8.69E-09 | RAB27A |
| 15 | 55868823 | G | A | 15:55868823_G/A | rs894868996 | 5.85 | 0.0001 | 0.0000 | 0.0023 | 68.1 | 11.8 | 7.38E-09 | PYGO1 |
| 17 | 60075778 | T | G | 17:60075778_T/G | rs945505625 | 22.53 | 0.0002 | 0.0002 | 0.0034 | 25.7 | 4.4 | 5.86E-09 | MED13 |
| 18 | 8103225 | G | A | 18:8103225_G/A | rs183665868 | 10.9 | 0.0001 | 0.0001 | 0.0022 | 46.7 | 8.2 | 1.01E-08 | PTPRM |

The presented variants were identified as genome-wide significant with $P \leq 5 \times 10^{-8}$ .

### Functional mapping and annotation of the non-HLA region

The SNP2GENE function prioritized three risk loci (2q35, 5p15.33, 6p21.2) from the summary statistics as the most likely to have a biological impact on CeD. The 2q35 and 6p21.2 loci mapped to nine protein coding genes. The lead SNP in 2q35 (rs189838725) was an intergenic variant mapping near four protein-coding genes: *IKZF2*, *SPAG16*, *VWC2L*, *ERBB4*. The lead SNP in 6p21.2 (rs192900921) was an intronic variant in *LRFN2* and mapping near four other protein-coding genes: *GLP1R*, *TSPO2*, *UNC5CL*, *APOBEC2*). As the three SNPs in the 5p15.33 locus (rs32723, rs32726, and lead SNP rs32727) were all located in a long non-coding RNA gene (*LINC01019*), the SNP2GENE function did not map *IRX1*, even though it was identified as the closest protein-coding gene in dbSNP.

The nine genes mapped to 2q35 and 6p21.2, were analyzed using the GENE2FUNC function to determine gene-tissue expressions and specificity via heatmap and DEG scores. Figure 4(a) shows the normalized expression of these genes across 54 tissues. *APOBEC2* was highly expressed in circulatory tissues, while *ERBB4*, *LRFN2*, *VWC2L*, and *SPAG16* were more expressed in neural tissues. *TSOP2* was expressed in whole blood, and *UNC5CL*, *IKZF2*, and *GLP1R* were expressed in renal, lymphatic, and digestive tissues. Figure 4(b) indicates tissues where the nine genes are up-regulated, down-regulated, or exhibit bi-directional expression (i.e. up-regulated in some tissues and down-regulated in other). Although none of the tissue regulations were significant and FUMA does not specify the direction of individual genes, they were generally up-regulated in the brain, pancreas, heart, and stomach, and down-regulated in the small intestine and esophagus. Bi-directional regulations were observed in gastrointestinal, nervous and circulatory tissues.

**Figure 4:**
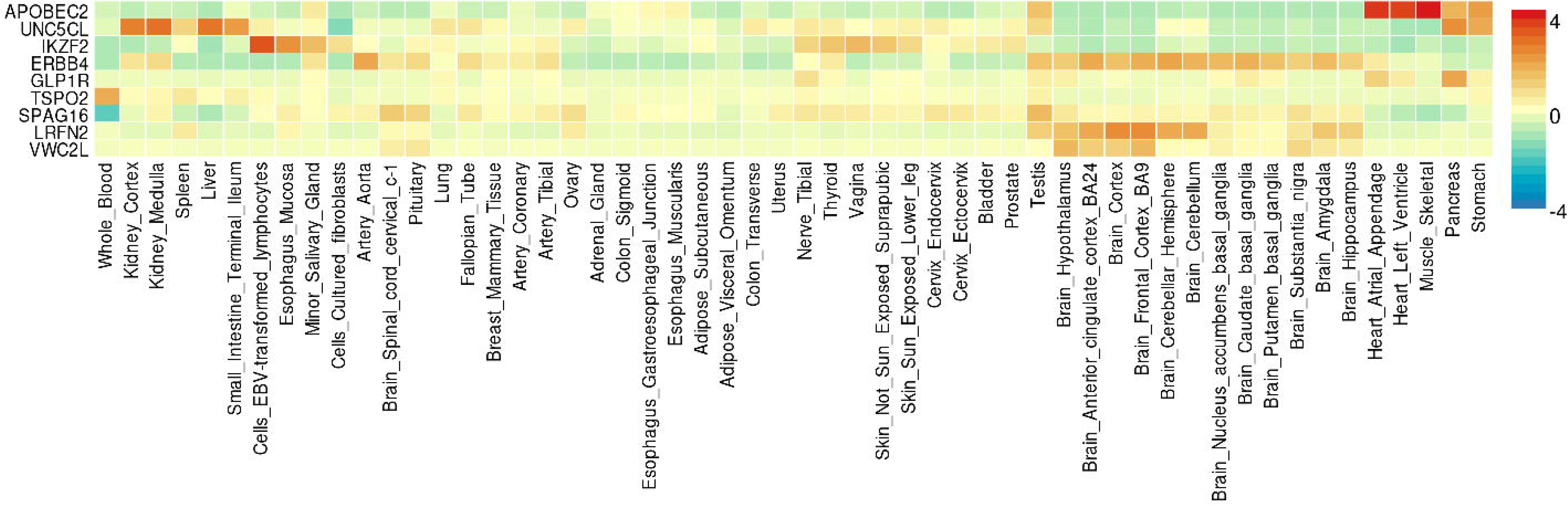
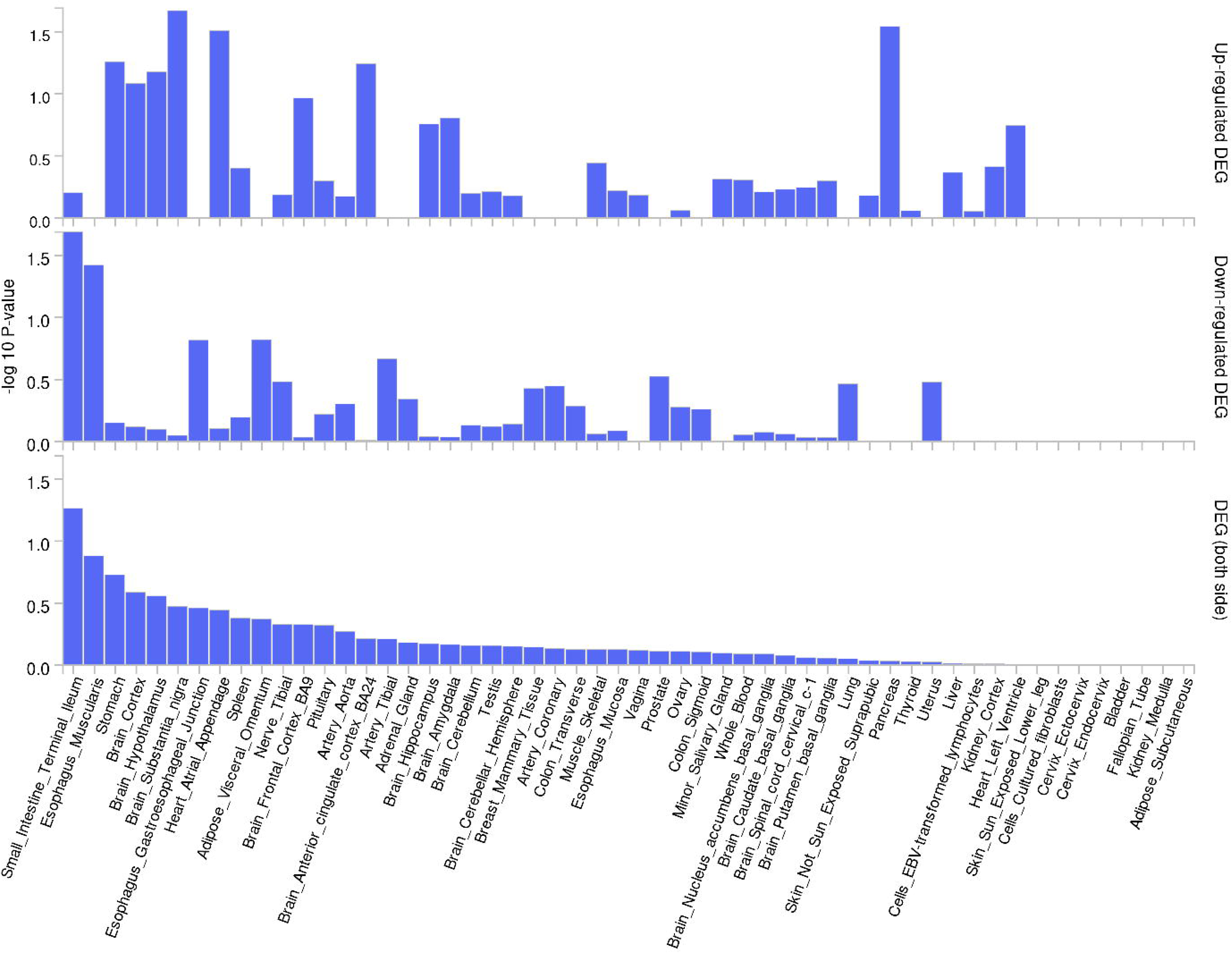
Gene Expression and Differentially Expressed Gene Enrichment from FUMA. **a) Gene Expression Heatmap Across 54 Human Tissues** The heatmap represents the normalized gene expression levels of nine genes (APOBEC2, UNC5CL, IKZF2, ERBB4, GLP1R, TSPO2, SPAG16, LRFN2, VWC2L) across 54 human tissues. The expression levels are scaled from -4 (blue) to +4 (red), with yellow representing median expression levels. The color scale on the right indicates the relative expression levels, where red corresponds to higher expression and blue indicates lower expression relative to the median. Each row represents a different gene, and each column corresponds to a specific tissue. **b) Differential Expression of Genes (DEGs) Across 54 Human Tissues** The bar plot displays the differential expression of nine genes (as shown in Figure 4a) across 54 human tissues. The x-axis lists the tissues, while the y-axis shows the -log10 P-values calculated by FUMA. Top panel: -log10 P-values for up-regulated DEGs in each tissue. Middle panel: -log10 P-values for down-regulated DEGs in each tissue. Bottom panel: Combined -log10 P-values for both up- and down-regulated DEGs, providing an overall view of significant DEGs across tissues. Higher bars indicate greater statistical significance (higher -log10 P-values), while shorter bars indicate lower significance.

### Genetic loci within the HLA-region associated with celiac disease

Three HLA loci were found to be significantly associated with CeD in the present population: two known loci (6p21.33 and 6p21.32) and one new locus (6p22.1) (Table 2). However, the poor coverage of the HLA region by the SNP array and imputation method used, led to non-representative findings in this region. Instead of the expected *HLA-DQB1* gene, the first lead SNP, rs2853999 (6:31326074_A/T; MAF >10% and P ≤ 3 × 10^−182^), located in the 6p21.33 locus, was identified as a two kilobases (KB) upstream variant of the *HLA-B* gene. The second lead SNP, rs715044 (6:29593788_G/T; MAF >5% and P ≤ 2.84 × 10^−32^) in the novel 6p22.1 locus, was identified as an intron variant within the *GABBR1* gene. After the third conditional analysis, rs28633132 (6:32626053_A/T; MAF >10% and P ≤ 4.97 × 10^−30^) emerged as the lead SNP. The variant is in the 6p21.32 locus, two kilobases upstream of the non-coding *HLA-DQB1-AS1* gene. Notably, the nearest protein-coding gene to this variant is *HLA-DQB1*. The stacked regional plot is illustrated in Figure 5.

**Figure 5:**
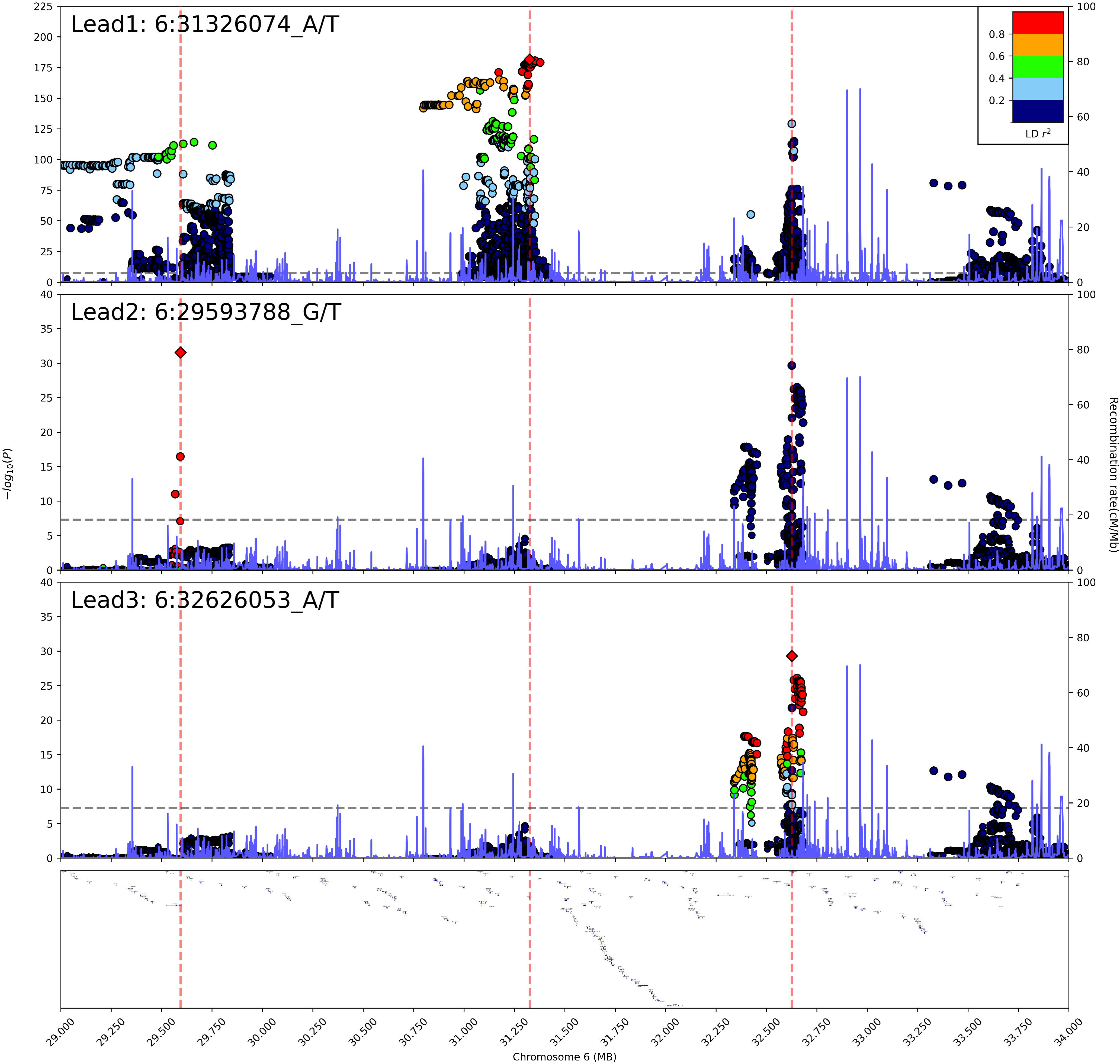
Stacked Regional Plot of the HLA-Region. Genetic variants are plotted according to chromosome and position (x-axis) and the-log_10_ P for the variant association (y-axis). The significance threshold (P ≤ 5 × 10^−8^) is indicated by the grey dashed line. The correlation r^2^ for each variant is indicated by colors relative to the index variants ±250 kilobase pairs. Three lead variants were observed in the HLA-region after conducting three conditional GWA analysis on all variants in LD (r2 >0.2): rs2853999, mapping to the *HLA-B gene*; rs715044, mapping to the novel *GABBR1* gene, and rs28633132, mapping to *HLA-DQB1-AS1,* a non-coding RNA with HLA-DQB1 as the closest protein-coding gene. HLA=human leukocyte antigen; GWA=Genome wide association; LD=Linkage disequilibrium.

**Table 2:** Lead variants in the HLA-region associated with celiac disease in the HUNT4 study.

| CHR | BP | REF | ALT | ID | RSID | MAC | MAF | AF<br>Cases | AF<br>Controls | BETA | SE | PVALUE | Genes |
| --- | --- | --- | --- | --- | --- | --- | --- | --- | --- | --- | --- | --- | --- |
| 6 | 31326074 | A | T | 6:31326074_A/T | rs2853999 | 13617 | 0.13 | 0.41 | 0.13 | 1.820 | 0.063 | 3.00E-182 | HLA-B |
| 6 | 29593788 | G | T | 6:29593788_G/T | rs715044 | 5369 | 0.05 | 0.05 | 0.05 | 1.524 | 0.129 | 2.84E-32 | GABBR1 |
| 6 | 32626053 | A | T | 6:32626053_A/T | rs28633132 | 11099 | 0.11 | 0.06 | 0.11 | -2.662 | 0.235 | 4.97E-30 | HLA-DQB1-AS1 |

The listed lead variants were identified through repeated regression analysis conditioned on all HLA-variants in LD ( $r^2$ $>0.2$ ). All associations have $P \leq 5 \times 10^{-8}$ . LD=Linkage disequilibrium.

**Table 3:**
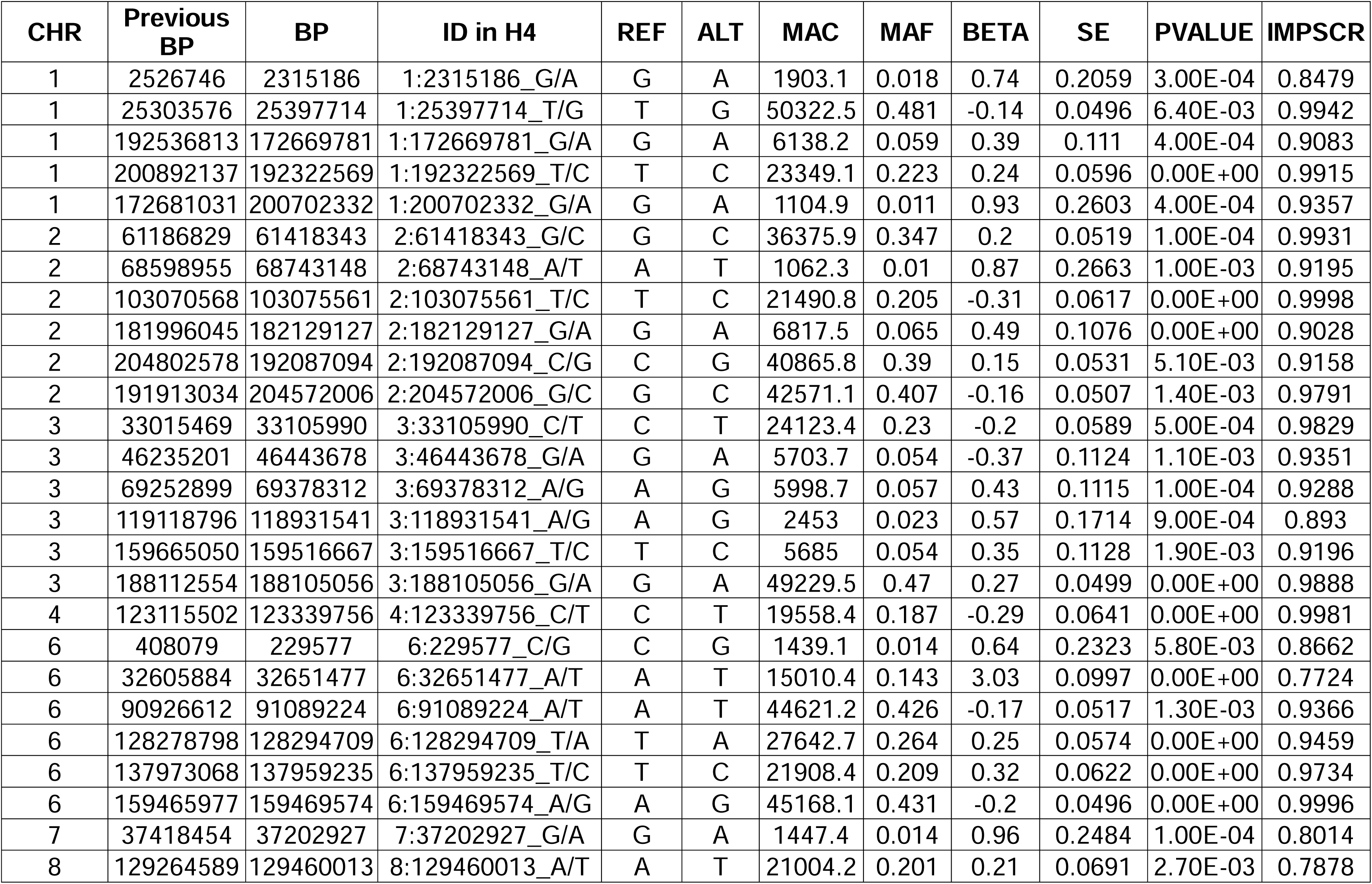
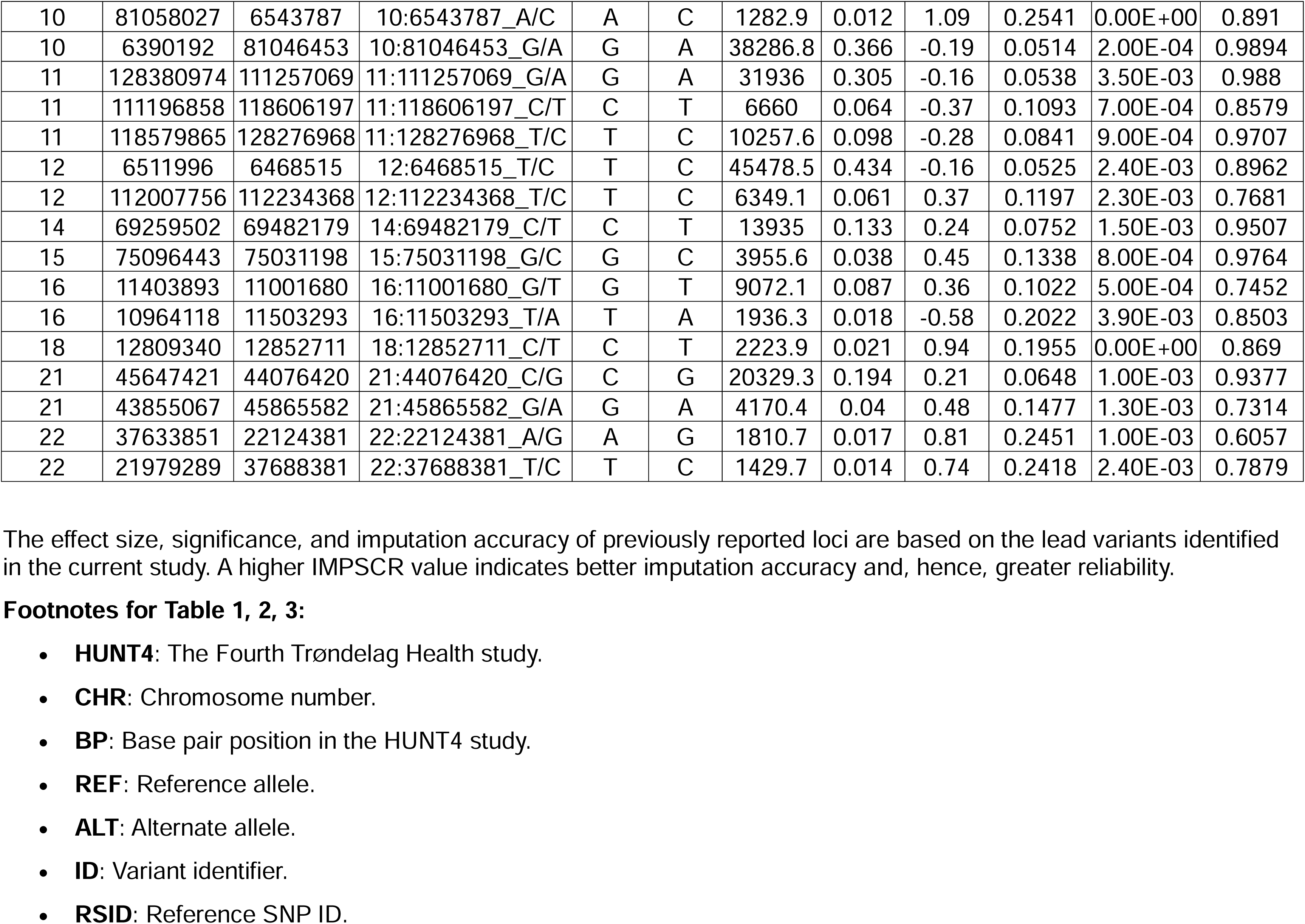
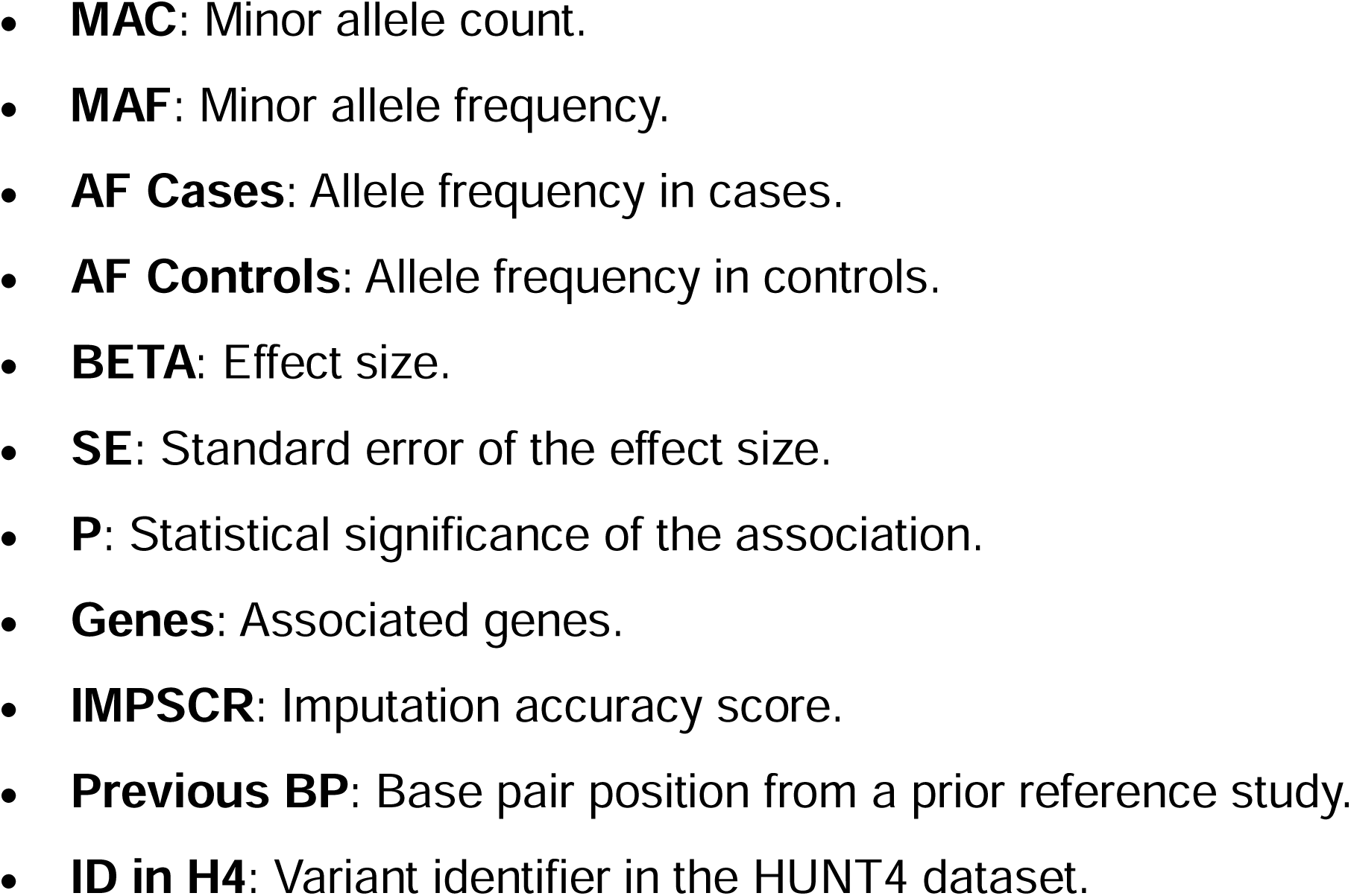
Lead variants and their corresponding imputation accuracy for known loci associated with celiac disease in the HUNT4 study.

### Heritability of celiac disease

The genome-wide non-HLA heritability (h^2^) on the liability scale for all confirmed CeD patients was estimated to be 23%±8%, using the LDSC function (36) in the GWASLab tool (37). The heritability increased from 7% for known cases to 11% for new cases, identified by the screening. The estimates were transformed from the observed to the liability scale based on the population prevalence rate of 1.5% reported in HUNT4 (23).

## Discussion

### Main findings

The present GWAS of a large adult population screened for CeD, including both previously known and unknown cases revealed 15 novel associations spread over 12 non-HLA loci. Out of these, the rs32727 SNP in the 5p15.33 locus was most promising. Additionally, the study identified 40 out of 41 previously known non-HLA loci. Among these six attained suggestive significances (P ≤ 5 × 10^−7^). The gene-tissue expression analysis suggested that the genes were mostly expressed in the small intestines, stomach, and brain. The gene expression in brain and blood-related tissues suggest a role of the genes in the identified loci outside the gut, which aligns with concurrent studies hypothesizing a gut-brain interaction in CeD (38,39).

### Novel associations

In support of a true association, the 5p15.33 locus consistently showed significant results in the subset analyses and was prioritized as the candidate with the strongest causal link to CeD in the functional analysis in FUMA. The lead SNP (rs32737) is mapped to a long non-coding RNA, complicating the interpretation of its clinical impact on CeD. Notably, the rs32737 variant is located close to the *IRX1* gene, which has been associated with rheumatoid arthritis (40), suggesting a new shared autoimmune locus. The *IRX1* gene, overexpressed in the bone marrow plasma cells, has been indicated to impact the production of the rheumatoid factor autoantibodies. Although the functional link with CeD is not clear, it has previously been shown that CeD shares risk loci outside the HLA-region with other autoimmune diseases, including rheumatoid arthritis and type 1 diabetes (41–43).

Among the remaining variants in novel loci, one was identified as non-protein coding (rs780153546), while the others mapped near genes involved in cellular transport (rs189838725, rs761616279), cellular structure and signaling (rs576626084, rs769377780, rs192900921, rs183665868, rs1002929661, rs137888770), or transcriptomic regulatory functions (rs551170288, rs945505625, rs894868996). In the only known locus, 2p16.1, the novel variant rs551170288 reached the GWAS significance threshold. This variant map downstream of the *BCL11A* gene, which encodes B-cell lymphoma 11A protein, often expressed in hematopoietic lineages and the brain. In CeD, the B-cells play a role in T-cell activation and subsequent villous blunting (44). Interestingly, the *BCL11A* gene houses in the same locus as *ASHA2* and *REL*, which are well established loci for CeD (15), with the latter involved in inflammation, immune response, and oncogenic processes. All novel variants should be interpreted with caution due to their low MAF and singleton association within the locus in our samples.

### Comparison to other GWAS findings

To our knowledge, this is the first GWAS conducted on a large adult population screened for CeD. It is also the first to implement a genotyping strategy emphasizing all genetic regions equally to identify novel variants associated with CeD. Consequently, the potentially novel 5p15.33 locus was not genotyped in the more recent GWASs (15,16). However, an earlier meta-analysis of 4,533 cases and 10,750 controls from 10 countries found no association between the 5p15.33 locus and CeD (13). The discrepancy with our results may be due to the different genotyping platforms used across cohorts and between cases and controls in the meta-analysis. Another factor could be a different allele frequency in the Norwegian population; however, this frequency was not reported in the meta-analysis (13). Our inclusion of undiagnosed CeD cases discovered through screening, who might have different symptoms or later onset than diagnosed cases (45), could hypothetically indicate a different genetic risk in this subgroup. Additionally, previous studies defined CeD diagnosis from hospital records or used less stringent diagnosis criteria (e.g. Marsh grade 1 or 2) (13–16). Moreover, while undiagnosed or potential cases were included in the control group of previous studies, diluting the association, the current study used a CeD-free control group. These factors may also explain why not all previous findings were replicated.

### Pathway Analysis

The pathway analysis prioritized three risk loci, of which 5p15.33 was in a non-coding region and thus not mapping to any protein-coding gene. The remaining loci, 2q35 and 6p21.2, mapped to nine genes. Both loci were rare variants with very low MAF and likely spurious associations. Nevertheless, these loci mapped to nine protein-coding genes, primarily expressed in the heart, stomach, muscle, kidney, liver, brain, and reproductive tissues. Although none of the tissue specificity estimates were significant, the brain, pancreas, heart, and stomach showed enhanced gene functionality, whereas gene functionality was suppressed in the small intestine, esophagus, and adipose tissues. The complex disease mechanisms are further suggested by the bi-directional gene regulation observed primarily in the small intestine, esophagus, and brain. The enrichment of gene expression in the small intestine and immune-related tissues is consistent with CeD development. Interestingly, the expression observed in brain and neural tissues supports the gut-brain interaction in CeD, as hypothesized in previous studies (38,39). The suggestive involvement of neural and other tissues could further help understand the extra-intestinal manifestation of CeD.

### HLA-related findings

The separate analysis of the HLA-region included a small number of loci, limiting us to detect novel associations in the HLA genes. The fact that the *HLA-DQB1* locus was not the most significantly associated with CeD in the current data suggests that the imputation does not accurately represent the true HLA region. This outcome was expected due to the reference panel used for imputation. Still, a novel intronic variant (rs715044) in the 6p22.1 locus, mapped to the *GABBR1* gene was strongly associated with CeD in this study. The 6p22.1 locus has also shown pleiotropic effects in rheumatoid arthritis and hypothyroidism (46) and is involved in encoding gamma-aminobutyric acid (GABA), the main inhibitory neurotransmitter in the nervous system. Interestingly, *GABBR1* is broadly expressed in the brain, which may relate to extraintestinal symptoms of CeD, such as brain fog and fatigue.

### Heritability

Notably, the current heritability estimates are lower than previous GWAS, 23% compared to 35% when calculated using the same method. This can be attributed to the different in genotyping and imputation methods used in these studies (13,15,47). While our screening strategy was stringent and high-quality SNP imputation was ensured, it lacked the focus on immune-specific loci that previous studies emphasized. Greater homogeneity in our cohort and the inclusion of all SNPs from the summary statistics file might have also affected the estimates. The difference in heritability for known and new cases may be attributed to differences in sample size, genetic makeup or environmental interaction effects. Due to the unclear cause of the loss of heritability, this estimate should be viewed with caution.

### Strengths and limitations

The major strength of this study is the screening of a population-based cohort with a comparatively high response rate, identifying both previously known, unknown, and potential CeD cases. This has minimized the selection and information biases observed in previous studies (13,15–17), arising from lower participation rates and including only known cases in the case group and unknown and potential cases in the control group. These biases may have attenuated previous associations with the 5p15.33 locus. Importantly, the SNP arrays and imputation panel used in the study were able to cover a wider range of non-HLA regions compared to arrays used earlier. In addition, the SAIGE tool enhanced the study by accounting for relatedness among the individuals and addressing imbalance in the number of cases and controls occurring when investigating a disease with a low prevalence in a large population. These strengths combined give more accurate and robust estimates compared to previous studies. A limitation is the lack of children and young adults from the screening. However, all adults were included regardless of their age at diagnosis, ensuring comprehensive coverage across all age groups. Nonetheless, some of the known cases were diagnosed many years ago when diagnostic methods were less precise, possibly leading to the inclusion of falsely diagnosed individuals in our study, which could attenuate potential genetic associations with CeD. Notably, this might also explain the higher heritability estimate for the new cases diagnosed with stringent criteria. Furthermore, it supports the idea that the screening reduced case misclassification, i.e., the information bias present in previous studies. Another limitation is the study’s restriction on the European population; hence the results may not be generalized for other ancestries. Finally, the small number of cases raise power concerns as it decreases the chances to detect rare genetic variants, especially potentially unique variants among the previously undiagnosed cases, which may represent a slightly different phenotype compared to cases identified through the health care system. It should also be noted that the HLA region is notoriously difficult to impute and interpret due to its large LD blocks, which can obscure associations with other causal SNPs. Additionally, the imputation panel used in the current study was not optimized for HLA analysis, leading to a considerable proportion of variants with low imputation values.

Ideally, a genotyping chip with a denser coverage in the whole genome, a combined imputation panel of HLA and non-HLA specific variants, and screening individuals of all age groups from different populations should be performed to be able to explain the larger part of the missing heritability in CeD. Additionally, analysis to determine the functional and causal pathways of the variants discovered is crucial to give a clearer clinical impact of the study’s result and its potential use in risk prediction and prevention strategies of CeD onset and prognosis.

## Conclusion

To conclude, this GWAS, conducted on a large general adult population screened for CeD, identified the novel 5p15.33 locus with the strongest association signal. The rs32727 variant, mapped to a long non-coding RNA region near the *IRX1* gene, which has been associated with rheumatoid arthritis, suggests a new shared locus for autoimmune disorders. However, further studies are warranted to replicate the association and validate a biological pathway between the variant and CeD to determine its potential clinical impact.

## Data Availability

All data produced in the present study are available upon reasonable request to HUNT Databank.

https://www.ntnu.edu/hunt/databank

## Acknowledgements

The Trøndelag Health Study (HUNT) is a collaboration between HUNT Research Centre (Faculty of Medicine and Health Sciences, Norwegian University of Science and Technology (NTNU) NTNU), Trøndelag County Council, Central Norway Regional Health Authority, and the Norwegian Institute of Public Health.

## Author contributions

Conceptualization: EN-J, RH

Data curation: MSA

Formal analysis: MSA

Funding acquisition: EN-J, RH, KH

Investigation: MSA

Methodology: MSA, LT, RH

Project administration: EN-J

Resources: EN-J

Software: MSA

Supervision: EN-J, RH, LMS, IHJ, SW, KEAL

Validation: MSA Visualization: MSA

Writing – original draft: MSA

Writing – review & editing: MSA, RH, EN-J, LMS, IHJ, LT, SW, KH, BB, KEAL

## Web resources

GWASLab - https://cloufield.github.io/gwaslab/

LDSC - https://cloufield.github.io/gwaslab/ldsc_in_gwaslab/

GeneCards - https://www.genecards.org/

dbSNP - https://www.ncbi.nlm.nih.gov/projects/SNP/get_html.cgi?whichHtml=overview

Opentargets - https://genetics.opentargets.org/

## Data and code availability

The Trøndelag Health Study (HUNT) has invited individuals aged 13-100 years to four surveys between 1984 and 2019. Comprehensive data from more than 140,000 individuals having participated at least once and biological material from 78,000 individual are collected. The data are stored in HUNT databank and biological material in HUNT biobank. HUNT Research Centre has permission from the Norwegian Data Inspectorate to store and handle these data. The key identification in the database is the personal identification number given to all Norwegians at birth or immigration, whilst de-identified data are sent to researchers upon approval of a research protocol by the Regional Ethical Committee and HUNT Research Centre. To protect participants’ privacy, HUNT Research Centre aims to limit storage of data outside HUNT databank and cannot deposit data in open repositories. HUNT databank has precise information on all data exported to different projects and are able to reproduce these on request. There are no restrictions regarding data export given approval of applications to HUNT Research Centre. For more information see: http://www.ntnu.edu/hunt/data

The codes supporting the current study have not been deposited in a public repository because the pipeline is under construction but are available from the corresponding author on request.

