## Supplementary figures and images for "Genome-wide association study identifies novel risk variants for celiac disease in the 5p15.33 locus: insights from a population-based screening of adults, the HUNT study"

### Supplementary file

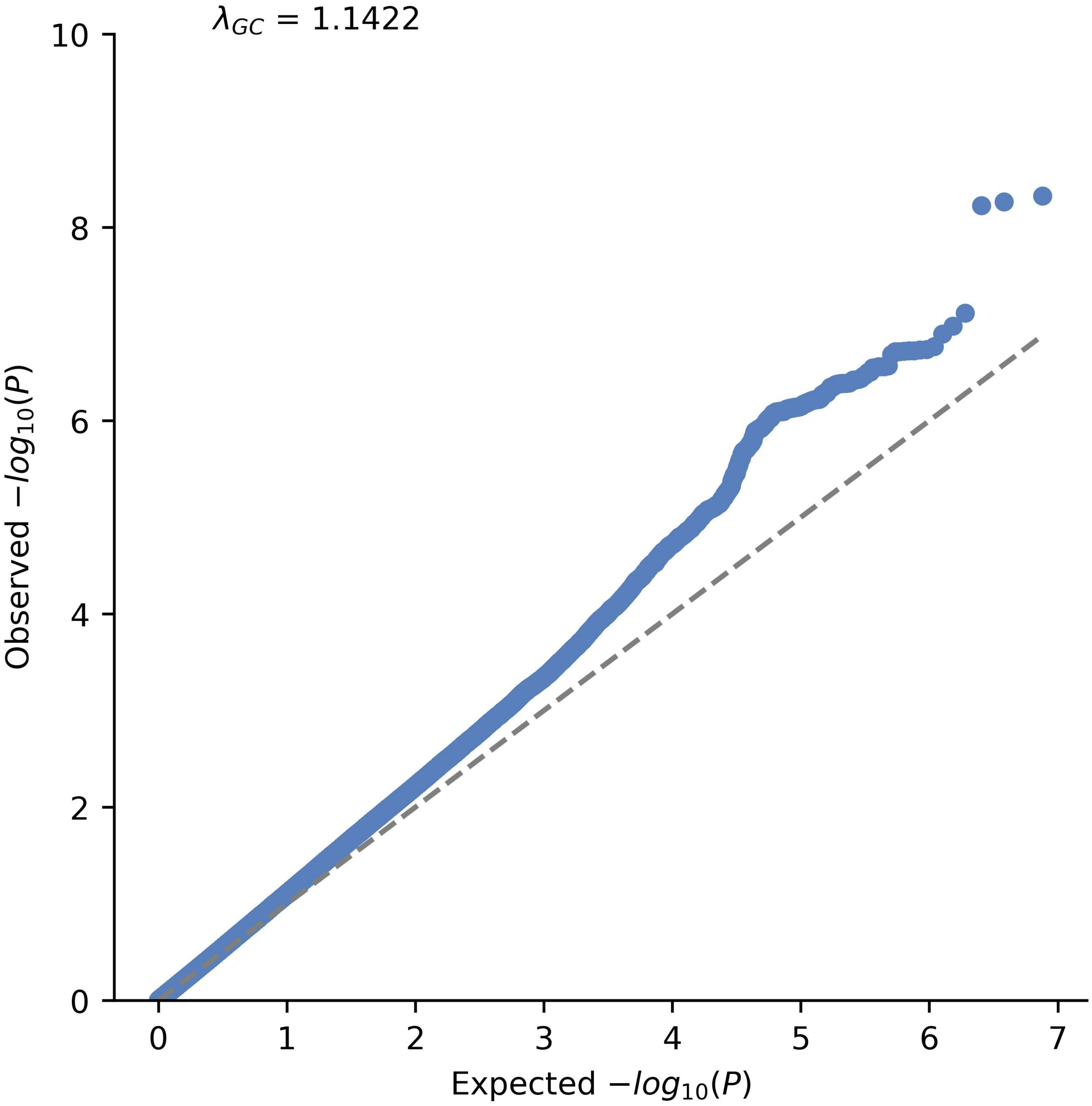

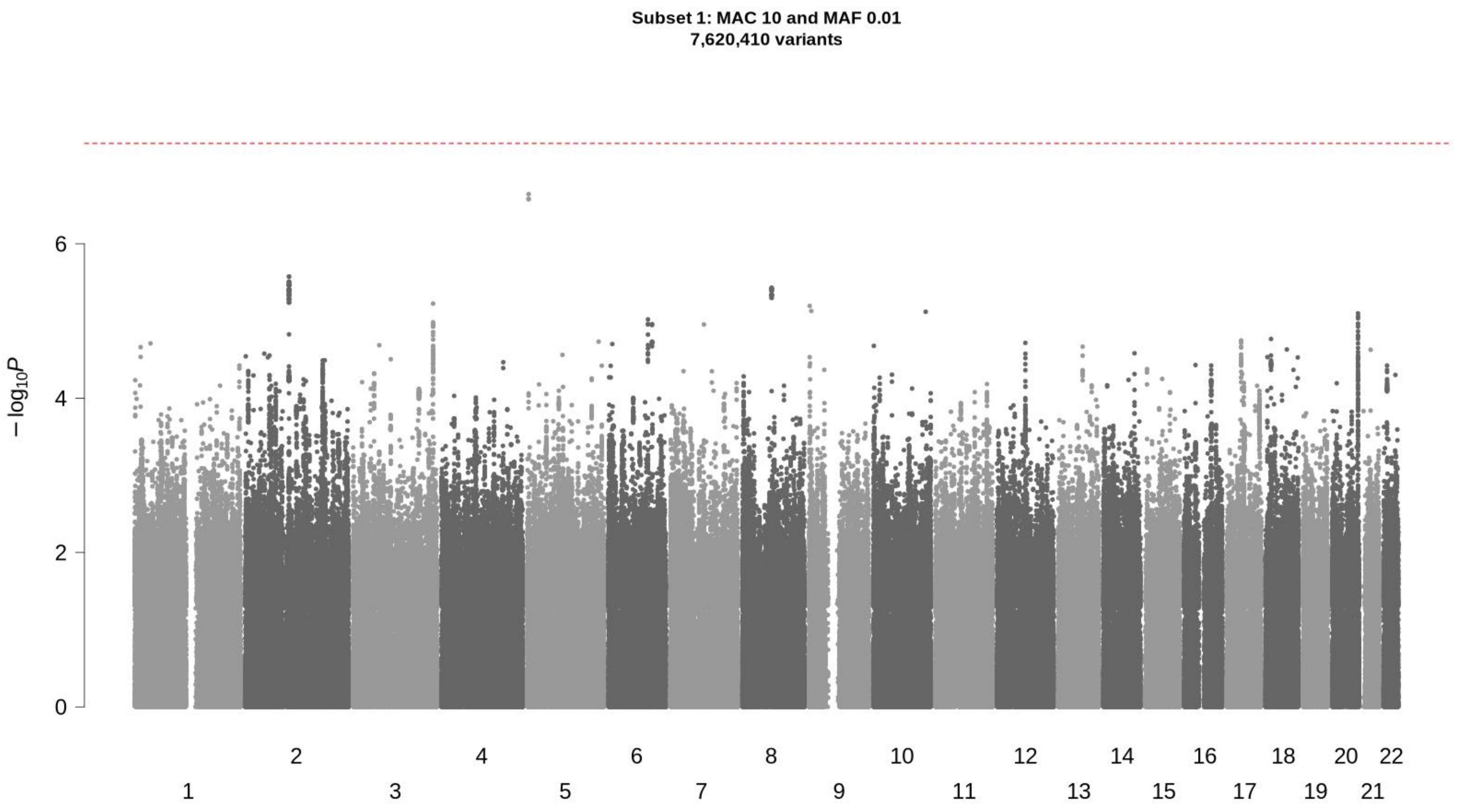

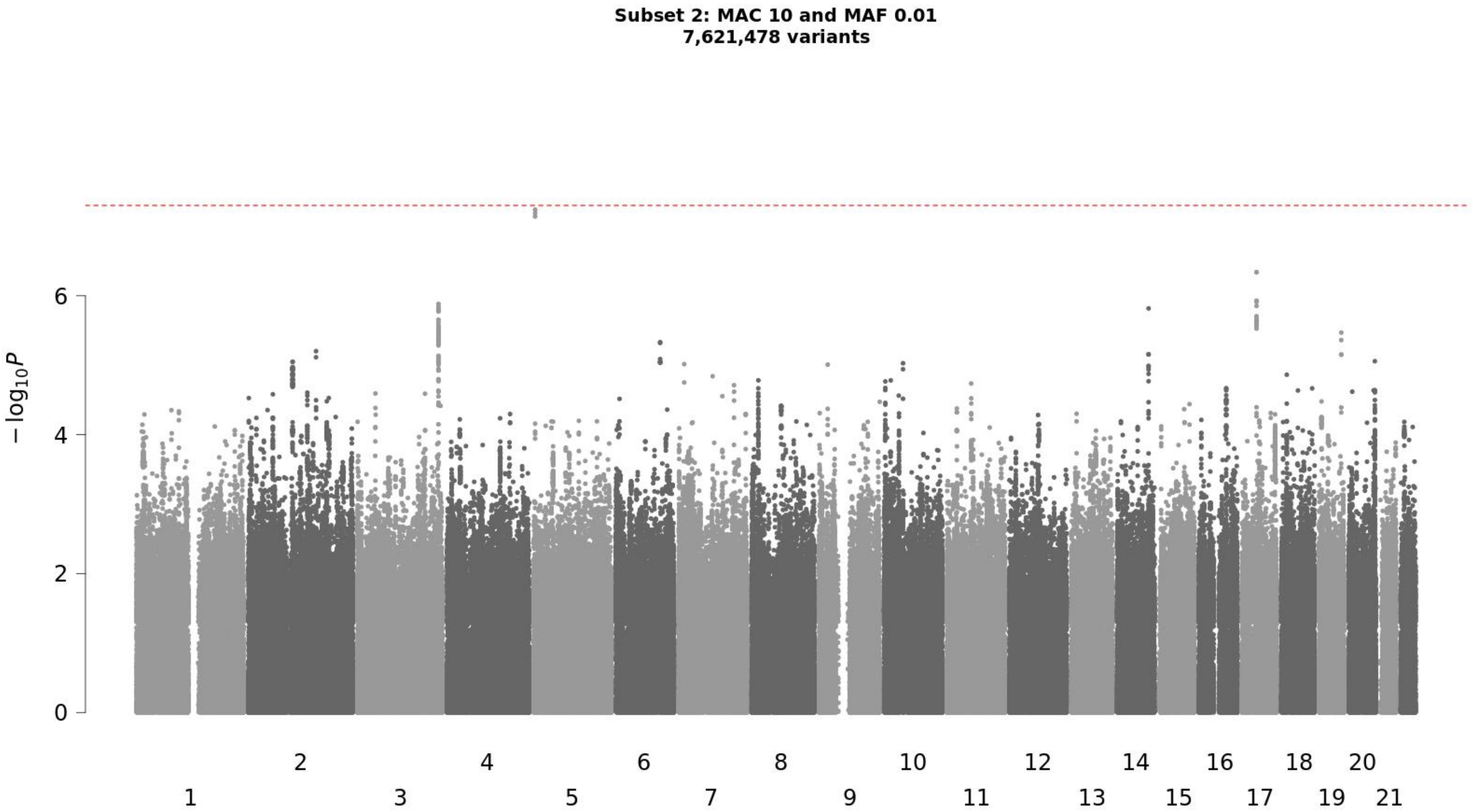

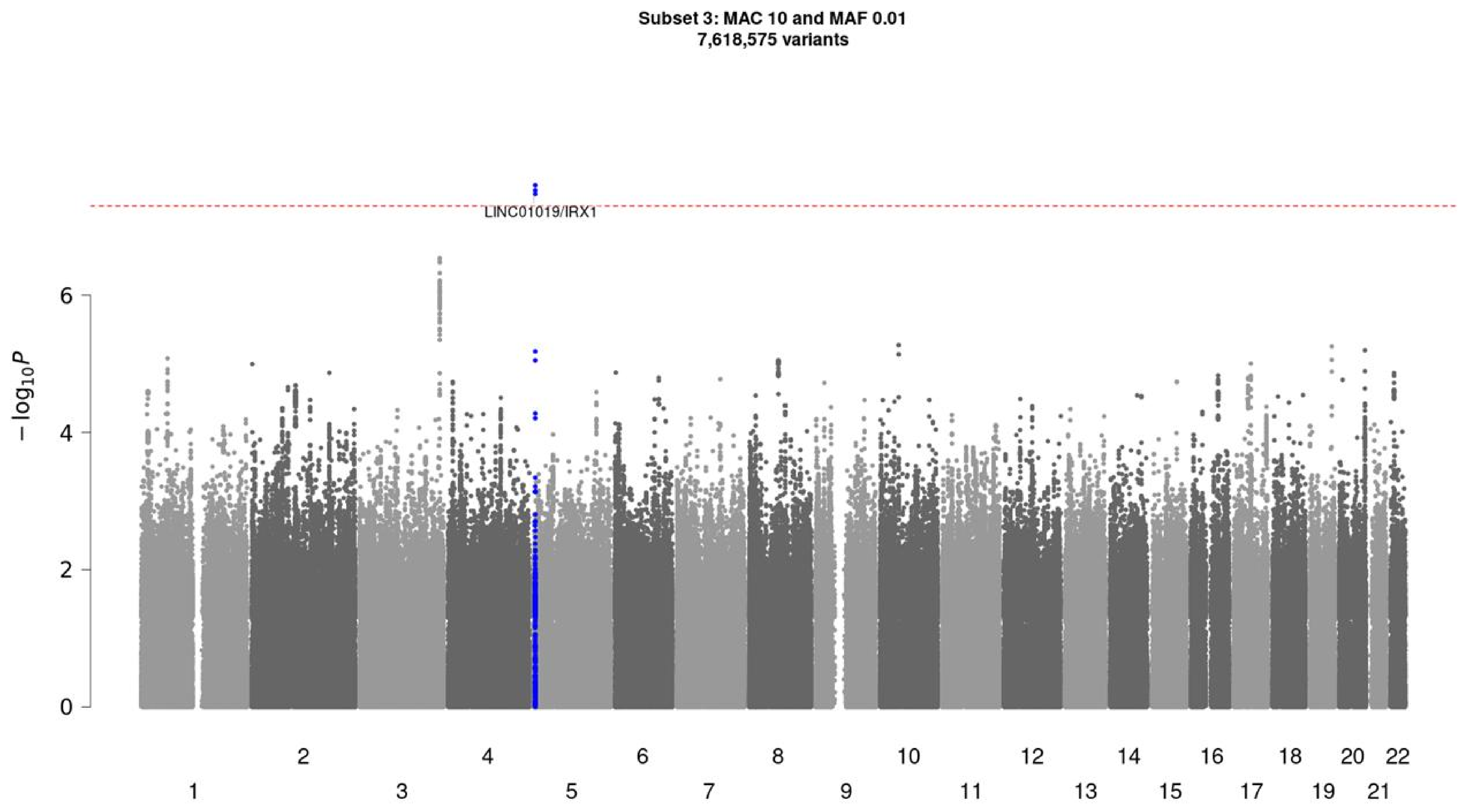

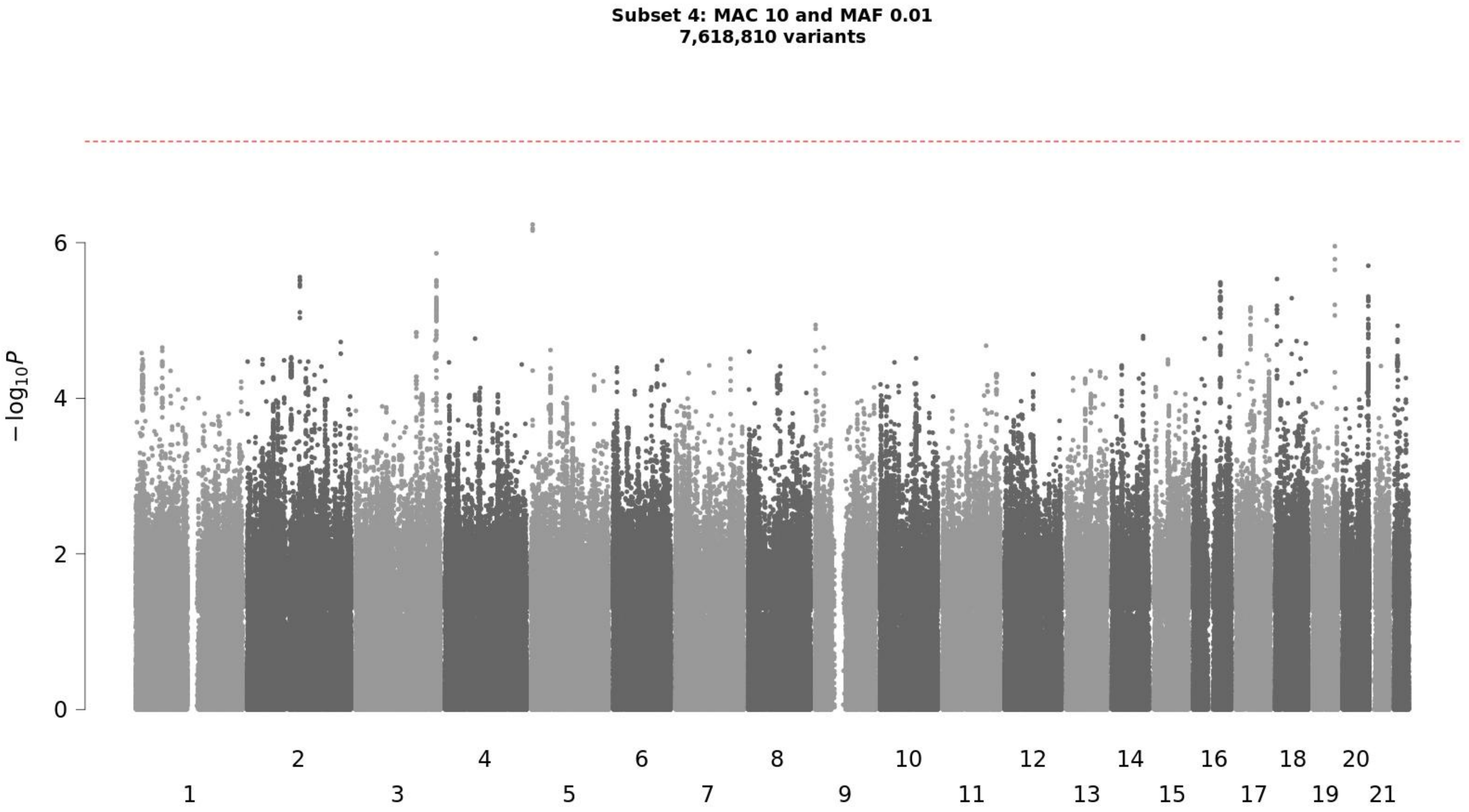

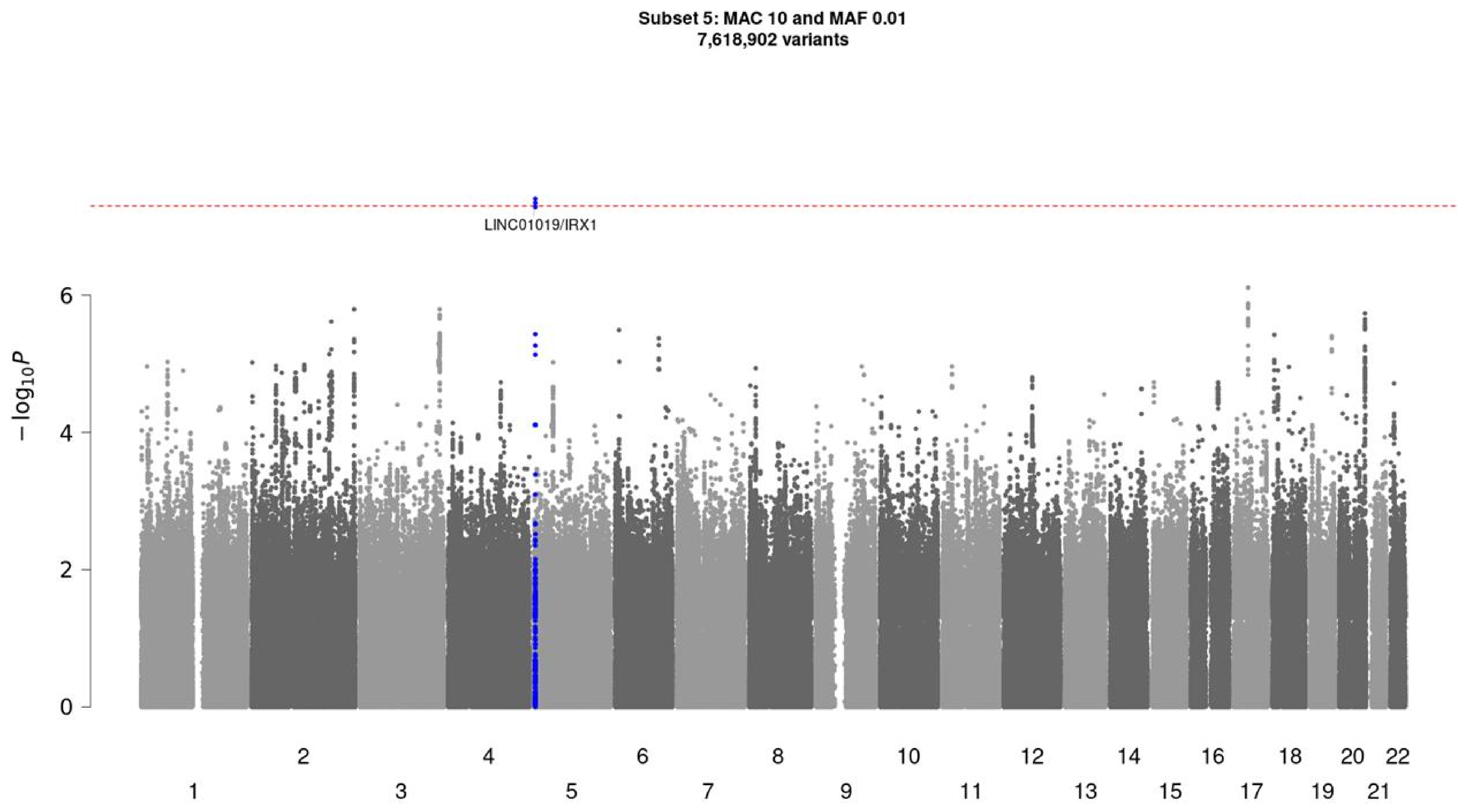

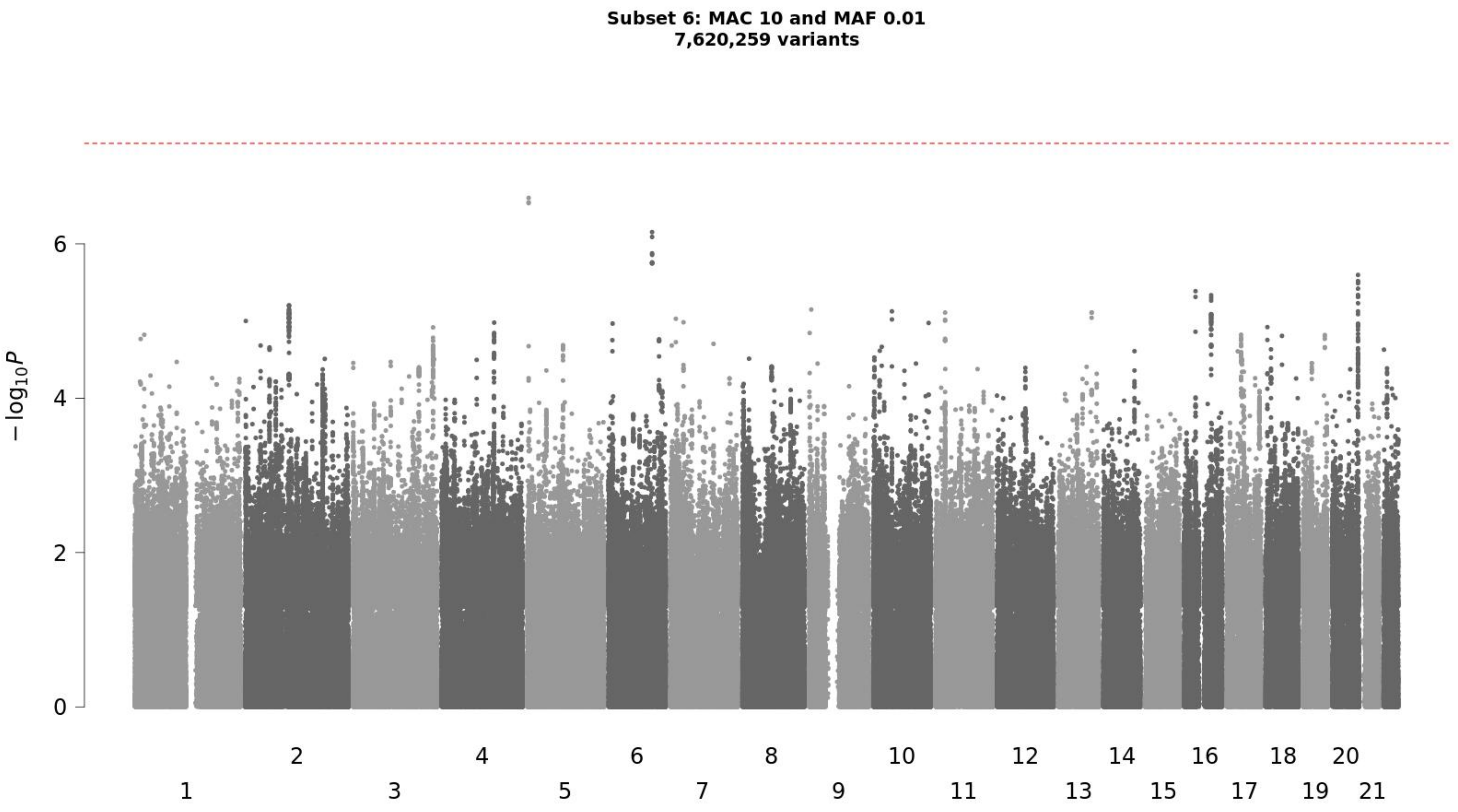
